# Occupational Physical Activity Paradox and Incident Cardiovascular Disease: Does Exposure Classification Matter?

**DOI:** 10.64898/2026.07.23.26358767

**Authors:** Winnifred Omono Okoro, Evangelia Demou, Carlos Celis-Morales, Gemma C. Ryde

**Affiliations:** School of Cardiovascular and Metabolic Health, University of Glasgow, Glasgow, UK, G12 8TA; School of Health and Wellbeing, University of Glasgow, Glasgow, UK, G12 8TB

**Keywords:** occupational physical activity, cardiovascular disease, cohort study, effect modification, occupational health, operationalisation, work exposure classification

## Abstract

**Objectives:** The association between occupational physical activity (OPA) and cardiovascular disease (CVD) may depend on how OPA is classified. This study examined the association between OPA and incident CVD using individual-level and occupational-group-level classifications, and assessed whether factors contributing to attenuation differed.

**Methods:** A total of 137,592 employed UK Biobank participants aged 40-69 years who were free of CVD at baseline were included. OPA was classified using (i) individual-level self-reported physical activity at work and (ii) occupational-group-level two-digit Standard Occupational Classification. Associations with incident CVD were examined using Cox proportional hazards models. Attenuation of associations by sociodemographic, lifestyle, health, occupational and psychosocial factors was assessed using univariate adjustments and Heller’s coefficient.

**Results:** During 1,656,378 person-years of follow-up (median 12.6 years), 10,465 incident CVD events occurred. The association between OPA and CVD differed markedly according to exposure classification. High individual-level self-reported OPA was associated with increased CVD incidence after full adjustment [hazard ratio (HR) 1.19, 95% confidence interval (Cl) 1.101.27]. In contrast, no association was observed when OPA was classified at the occupational-group level using Standard Occupational Classification groupings (HR 1.00, 95% CI 0.94-1.06). Sociodemographic and health factors accounted for the greatest attenuation across both classifications, whereas psychosocial factors contributed primarily to associations based on individual-level self-reported OPA.

**Conclusions:** The association between occupational physical activity and incident cardiovascular disease differed substantially according to how OPA was classified. These findings highlight exposure classification as a critical consideration when interpreting evidence on the physical activity paradox.

**What is already known on this topic?:**

- The “physical activity paradox” suggests that high occupational physical activity may increase cardiovascular disease risk, in contrast to the established benefits of leisure-time physical activity.
- Evidence on the health effects of occupational physical activity is inconsistent, with heterogeneity in how occupational physical activity is operationalised potentially contributing to divergent findings.
- No previous UK Biobank study has examined the association between occupational physical activity and incident cardiovascular disease while comparing alternative approaches to occupational physical activity operationalisation.

**What this study adds?:**

- This study provides the first evidence from the UK Biobank on the association between occupational physical activity and incident cardiovascular disease, showing increased risk at higher levels of individual-level self-reported occupational physical activity.
- Findings differed substantially according to how occupational physical activity was classified: individual-level self-reported occupational physical activity was associated with increased cardiovascular disease risk, whereas a group-based Standard Occupational Classification approach showed no association, demonstrating that exposure classification meaningfully influences conclusions about the occupational physical activity-cardiovascular disease association.
- Socioeconomic and health factors accounted for the greatest attenuation across both approaches, while psychosocial factors contributed primarily to associations based on individual-level self-reported occupational physical activity, highlighting that the explanatory factors most apparent in occupational physical activity - cardiovascular disease associations may also depend on how occupational physical activity is classified.

**How this study might affect research, practice and policy:**

- The findings suggest that future research should explicitly consider how occupational physical activity is operationalised and avoid assuming that individual-level and group-level classifications are interchangeable. Occupational health research and guidance should interpret occupational physical activity-related cardiovascular risk in the context of the exposure classification used and consider the broader social and health characteristics of workers.

## INTRODUCTION

Physical activity (PA) is widely promoted as a cornerstone of cardiovascular disease (CVD) prevention, and current guidelines generally present PA as beneficial regardless of the domain in which it is performed [1, 2], However, the “physical activity paradox” challenges this assumption by suggesting that occupational physical activity (OPA), unlike leisure-time PA, may be associated with adverse cardiovascular health outcomes [3, 4], This question is particularly important given that CVD remains a major public health concern globally, affecting more than 7.6 million people in the United Kingdom (UK) alone, and accounting for approximately 26% of deaths [5]. As work represents a substantial component of daily activity for many adults, understanding whether OPA is associated with cardiovascular risk is essential for interpreting the health effects of occupational exposures and for informing appropriately nuanced PA guidance [6].

Despite considerable research, evidence on the association between OPA and CVD and the PA paradox remains inconsistent across studies, populations and outcomes [7]. An umbrella review of 158 studies reported favourable associations between OPA and stroke and coronary heart disease, higher all-cause mortality among men and no association with allcause mortality among women [3]. Similarly, a meta-analysis of 103 prospective cohort studies found no protective association between OPA and total CVD, coronary heart disease, stroke, or atrial fibrillation [8]. Findings also differ between countries with broadly comparable socioeconomic and occupational contexts. For example, Norwegian cohort data have suggested longer life expectancy among individuals in physically demanding occupations [9], whereas Danish data have linked high OPA with increased risks of CVD and all-cause mortality [10]. These inconsistencies suggest that the relationship between OPA and CVD is nuanced and may be shaped by a range of factors.

Before further interpreting the factors contributing to this relationship, it is necessary to establish whether observed differences may partly reflect how OPA is defined and classified. OPA has been operationalised in markedly different ways across studies, including individual self-reported measures and occupational-group approaches such as job exposure matrices (JEMs) or classifications based on job title [3, 4], Individual-level measures reflect each worker’s reported physical demands, whereas occupational-group approaches assign the same exposure to everyone within an occupational group. Consequently, the same worker may be classified differently and different dimensions of occupational exposure captured.

Occupational epidemiological research has recognised that individual- and occupational-group-level exposure assessments involve distinct trade-offs between exposure specificity and feasibility and may therefore produce different estimates of exposure-health associations [11, 12]. Despite this, few studies have systematically examined whether the association between OPA and incident CVD differs according to the level at which OPA is classified, particularly in relation to the PA paradox.

In addition, evidence on whether associations differ across worker groups is also limited [3]. Existing findings suggest that associations may differ across sociodemographic groups, with high OPA reported as protective among lower-educated workers in China [13] and younger workers in Sweden [14], but more harmful among lower-income groups in United States cohorts [15]. Psychosocial characteristics, including job satisfaction and happiness, which are themselves associated with chronic disease risk [16], are rarely considered. As a result, potentially vulnerable worker subgroups may remain obscured within aggregate estimates of OPA-related risk.

This study investigates the association between OPAand incident CVD using individual-level self-reported OPA and group-level occupational classifications to assess the extent to which observed associations depend on how OPA is operationalized. Incident CVD was selected as the primary outcome as it reflects disease onset rather than mortality which may also be influenced by survival after diagnosis and competing causes of death. We further examine whether observed associations are robust to sociodemographic, lifestyle, health, occupational and psychosocial factors and explore potential heterogeneity across relevant worker subgroups. By directly comparing alternative OPA classifications, this study aims to clarify whether inconsistencies in the literature regarding the PA paradox reflect genuine differences in occupational cardiovascular risk or, in part, differences in exposure definition.

## METHODS

### Study Design and Population

This study used data from the UK Biobank, a large prospective cohort of over 500,000 adults aged 40-69 years, recruited between 2006 and 2010 through National Health Service (NHS) general practitioner registries across England, Scotland, and Wales [17]. At baseline, participants completed self-administered touch-screen questionnaires, computer-assisted interviews, and physical assessments capturing sociodemographic characteristics (including age, sex, ethnicity, educational attainment, household income, and area-level deprivation), lifestyle behaviors (smoking status, alcohol intake, and diet quality), occupational characteristics (shift work, weekly working hours, and employment duration), psychosocial factors (job satisfaction and happiness), and health-related measures (body mass index, blood pressure, and physician-diagnosed conditions). Health outcomes were ascertained through linkage to inpatient hospital admission records and national death registries [18]. The UK Biobank study received ethical approval from the Northwest Multi-Centre Research Ethics Committee (reference ll/NW/0382 - Biobank project number 71392), and all participants provided written informed consent.

Between 2013 and 2015, a subsample of participants (approximately 100,000) wore a wrist-mounted triaxial accelerometer (Axivity AX3) continuously for seven days [18]. Raw acceleration data were processed using established UK Biobank protocols to derive estimates of sedentary behavior and moderate-to-vigorous physical activity (MVPA)[19]. These measures were used descriptively to characterize baseline PA profiles across OPA categories.

For the present analysis, participants were included if they were employed at baseline and had complete self-reported OPA data. Participants with missing exposure or covariate data or prevalent CVD at baseline were excluded, resulting in a final analytical sample of 137,592 participants. Participant exclusions are detailed in Figure 1.

**Figure 1:**
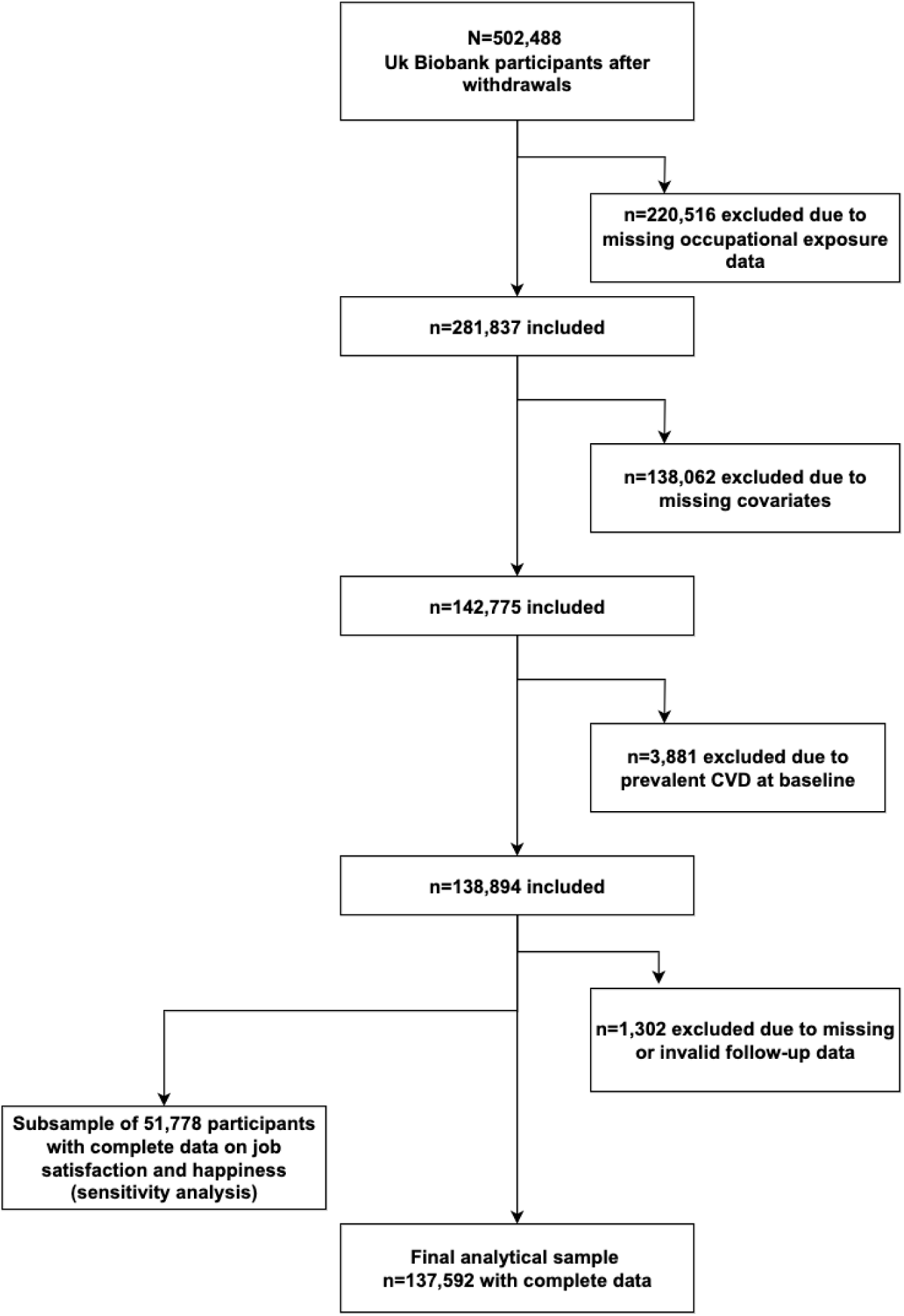
Flow Diagram of Participant Inclusion and Exclusion Criteria.

### Occupational Physical Activity Exposures

OPA was assessed using two complementary approaches: (i) an individual-level classification based on participants’ self-reported PA at work and (ii) an occupational-group-level classification based on Standard Occupational Classification (SOC) codes. The latter used a JEM informed approach, assigning each occupational group a single OPA category according to the predominant distribution of self-reported OPA within that occupation. Consequently, the individual-level approach assigned exposure according to each participant’s own reported physical demands, whereas the occupational-group-level approach assigned exposure according to the typical physical demands of the occupational group, irrespective of individual variation within that occupation.

### Individual-level OPA Classification

Individual-level OPA was assessed with self-reported OPA using two questions: “Does your job involve heavy manual or physical work?” and “Does your job involve standing or walking?”. Each item was answered on a four-point scale (“never/rarely”, “sometimes”, “usually”, “always퀝) and collapsed into three levels: low, moderate, and high. These two domains were cross tabulated to generate nine mutually exclusive exposure groups representing all combinations of manual/physical work and standing/walking demands. For primary analyses, these groups were collapsed into four composite OPA categories (very light, light, moderate, and high) to improve interpretability and statistical power while preserving exposure contrast. Full details of the nine exposure groups are provided in Supplementary Table S1.

### Group-Level OPA Classification

Group-level OPA was assessed with group-level OPA using participants’ two-digit Standard Occupational Classification (SOC) codes (sub-major group level). Each SOC group was assigned to one of three occupational activity categories: inactive, mixed, or physically demanding, based on the predominant physical demands of self-reported OPA within each occupational group. SOC groups in which ≥70% of participants reported very light or light OPA were classified as inactive, whereas those in which ≥60% reported moderate or high OPA were classified as physically demanding. SOC groups with a more even distribution across OPA categories were classified as mixed. This dominance-based approach is consistent with previous occupational epidemiology studies that classify occupations according to predominant physical demands of that occupation rather than their individual reported activity level [4, 9, 20–22] Examples of occupations within each category are provided in Supplementary Table S2.

### Outcomes

The primary outcome was incident CVD, defined using ICD-10 codes 100-199. Participants were followed from baseline until first event, death, loss to follow-up or end of available follow-up.

### Covariates

Potential confounders were selected based on covariates commonly reported in existing literature[7, 8]. *Sociodemographic factors* included age (years), sex (male, female), ethnicity (White, Mixed, South Asian, Black, Chinese), educational attainment (no qualification, other qualification, college/university degree), household income (<£18,000, £18,000-£31,000, £31,000-<£52,000, £52,000-£100,000, >£100,000), and Townsend deprivation index (low, middle, high deprivation).

*Lifestyle factors* included smoking status (never, former, current), alcohol intake (units per week), and diet quality (most healthy, moderately healthy, less healthy). Leisure-time PA was approximated using the UK Biobank “type of PA” variable, which captures engagement in up to eight activity categories (none, walking, exercise, sports, DIY, two types, three types, four types, or all types) and was used as a proxy measure of leisure-time activity.

*Occupational factors* included shift work (never/rarely, sometimes, usually, always), weekly working hours (<35, 35-40, >40 hours), and years in employment (1-9, 10-19, ≥20 years). *Health-related factors* included body mass index (continuous), systolic blood pressure (continuous), physician-diagnosed diabetes (yes/no), physician-diagnosed hypertension (yes/no), and multimorbidity (none, one condition, two or more conditions). *Psychosocial factors* included job satisfaction (extremely satisfied to extremely dissatisfied) and happiness (extremely happy to extremely unhappy).

### Statistical analyses

Baseline characteristics were summarized across OPA categories using means and standard deviations for continuous variables and frequencies with percentages for categorical variables. Cox proportional hazards regression models were used to estimate hazard ratios (HRs) and 95% confidence intervals (Cis) for associations between OPA categories and incident cardiovascular disease (CVD).

A sequential modelling strategy was employed. Model 0 was unadjusted. Model 1 adjusted for age and sex. Model 2 further adjusted for sociodemographic, lifestyle, and health-related covariates. Model 3 additionally adjusted for occupational factors, including shift work, years in employment, and weekly working hours. Model 4 further adjusted for type of PA, used as a proxy for leisure-time PA. A sensitivity model additionally included psychosocial factors (job satisfaction and happiness).

Sex-stratified analyses were conducted to examine potential differences in associations between men and women. For secondary analyses, effect modification of the OPA-CVD association was assessed within the high OPA group and within physically demanding occupations using univariate regression models. Each covariate was sequentially introduced into the unadjusted model, and Heller’s coefficient was used to quantify the proportion of risk attenuation attributable to individual factors.

Statistical significance was defined as a two-sided p<0.05. All analyses were conducted using Stata version 18.0 MP (StataCorp, College Station, TX), and results were visualized using GraphPad Prism version 10 (GraphPad Software, San Diego, CA).

## RESULTS

### Baseline Characteristics and Occupational Profiles

The final analytical sample comprised 137,592 participants with a mean age of 52.0 ± 7.0 years. Baseline characteristics by OPA category are presented in Table 1.

**Table 1.** Baseline Characteristics of Participants Across Occupational Physical Activity (OPA) Exposure Groups.

| Mean (SD) / n (%) | Very Light<br>(N=49,767) | Light<br>(N=43,574) | Moderate<br>(N=29,517) | Highest (N=14,734) |
| --- | --- | --- | --- | --- |
| **Age, years** | 52.0 (7.0) | 52.7 (7.0) | 52.9 (7.0) | 52.0 (7.1) |
| **Sex** |  |  |  |  |
| Females | 27,382 (55.0%) | 21,914 (50.3%) | 16,701 (56.6%) | 5,558 (37.7%) |
| Males | 22,385 (45.0%) | 21,660 (49.7%) | 12,816 (43.4%) | 9,176 (62.3%) |
| **Townsend deprivation index** |  |  |  |  |
| Lower deprivation | 18,123 (36.4%) | 15,483 (35.5%) | 9,637 (32.6%) | 3,885 (26.4%) |
| Middle | 16,912 (34.0%) | 15,269 (35.0%) | 10,107 (34.2%) | 5,000 (33.9%) |
| Higher deprivation | 14,732 (29.6%) | 12,822 (29.4%) | 9,773 (33.1%) | 5,849 (39.7%) |
| **Household Income** |  |  |  |  |
| <£18,000 | 2,397 (4.8%) | 2,931 (6.7%) | 3,967 (13.4%) | 2,997 (20.3%) |
| £18,000 -30,999 | 7,584 (15.2%) | 8,126 (18.6%) | 7,487 (25.4%) | 5,088 (34.5%) |
| £31,000 to 51,999 | 14,642 (29.4%) | 14,269 (32.7%) | 9,888 (33.5%) | 4,628 (31.4%) |
| £ 52,000 to 100,000 | 18,642 (37.5%) | 14,605 (33.5%) | 7,081 (24.0%) | 1,809 (12.3%) |
| >£ 100,000 | 6,502 (13.1%) | 3,643 (8.4%) | 1,094 (3.7%) | 212 (1.4%) |
| **Educational Qualification** |  |  |  |  |
| No qualification | 1,893 (3.8%) | 3,108 (7.1%) | 4,226 (14.3%) | 3,947 (26.8%) |
| Other qualification | 21,639 (43.5%) | 19,582 (44.9%) | 13,685 (46.4%) | 8,456 (57.4%) |
| College/University degree | 26,235 (52.7%) | 20,884 (47.9%) | 11,606 (39.3%) | 2,331 (15.8%) |
| **Shift work** |  |  |  |  |
| Never/rarely | 46,932 (94.3%) | 37,466 (86.0%) | 22,465 (76.1%) | 9,555 (64.9%) |
| Sometimes | 1,433 (2.9%) | 3,454 (7.9%) | 2,692 (9.1%) | 1,776 (12.1%) |
| Usually | 324 (0.7%) | 714 (1.6%) | 852 (2.9%) | 606 (4.1%) |
| Always | 1,078 (2.2%) | 1,940 (4.5%) | 3,508 (11.9%) | 2,797 (19.0%) |
| **Years in employment** |  |  |  |  |
| 1–9 years | 23,783 (47.8%) | 19,353 (44.4%) | 13,188 (44.7%) | 6,520 (44.3%) |
| 10–19 years | 13,284 (26.7%) | 11,814 (27.1%) | 7,793 (26.4%) | 3,549 (24.1%) |
| 20+ years | 12,700 (25.5%) | 12,407 (28.5%) | 8,536 (28.9%) | 4,665 (31.7%) |
| **Weekly working hours** |  |  |  |  |
| <35 hrs | 15,306 (30.8%) | 12,852 (29.5%) | 11,903 (40.3%) | 4,587 (31.1%) |
| 35–40 hrs | 21,448 (43.1%) | 17,084 (39.2%) | 9,896 (33.5%) | 5,517 (37.4%) |
| >40 hrs | 13,013 (26.1%) | 13,638 (31.3%) | 7,718 (26.1%) | 4,630 (31.4%) |
| **Smoking status** |  |  |  |  |
| Never | 29,735 (59.7%) | 25,422 (58.3%) | 17,128 (58.0%) | 7,758 (52.7%) |
| Previous | 15,983 (32.1%) | 14,297 (32.8%) | 9,235 (31.3%) | 4,753 (32.3%) |
| Current | 4,049 (8.1%) | 3,855 (8.8%) | 3,154 (10.7%) | 2,223 (15.1%) |
| **Alcohol (Units/week)** | 16.61 (16.61) | 16.97 (17.46) | 16.41 (19.04) | 20.73 (23.42) |
| **Diet quality** |  |  |  |  |
| Most Healthy | 25,974 (52.2%) | 22,987 (52.8%) | 15,140 (51.3%) | 6,194 (42.0%) |
| Moderately healthy | 11,223 (22.6%) | 9,647 (22.1%) | 6,276 (21.3%) | 3,348 (22.7%) |
| Less healthy | 12,570 (25.3%) | 10,940 (25.1%) | 8,101 (27.4%) | 5,192 (35.2%) |
| **BMI categories** |  |  |  |  |
| Underweight | 257 (0.5%) | 220 (0.5%) | 148 (0.5%) | 54 (0.4%) |
| Normal | 19,354 (38.9%) | 14,964 (34.3%) | 10,466 (35.5%) | 4,700 (31.9%) |
| Overweight | 20,401 (41.0%) | 18,767 (43.1%) | 12,561 (42.6%) | 6,592 (44.7%) |
| Obese | 9,755 (19.6%) | 9,623 (22.1%) | 6,342 (21.5%) | 3,388 (23.0%) |
| **Systolic Blood Pressure** | 133.38 (17.31) | 134.68 (17.62) | 135.47 (17.88) | 137.33 (17.71) |
| **Accelerometer sedentary time (min.day)** | 1076.4 (78.2) | 1065.7 (78.6) | 1047.8 (83.7) | 1026.2 (96.2) |
| **Accelerometer Light PA (min.day)** | 286.0 (59.1) | 296.1 (58.9) | 310.0 (61.6) | 319.2 (66.4) |
| **Accelerometer Moderate PA (min.day)** | 72.2 (31.1) | 73.0 (31.6) | 76.8 (34.7) | 88.3 (43.6) |
| **Accelerometer vigorous PA (min.day)** | 5.4 (6.8) | 5.2 (6.4) | 5.4 (6.6) | 6.3 (6.9) |
| **Accelerometer MVPA (met.min.week)** | 2323.2 (1078.4) | 2335.8 (1079.7) | 2451.0 (1179.1) | 2823.7 (1451.5) |
| **Fitness (Vo2max_mlkgmin)** | 35.8 (9.2) | 35.4 (9.1) | 34.3 (9.0) | 35.9 (9.3) |
| **Multimorbidity** |  |  |  |  |
| None | 22,785 (45.8%) | 19,205 (44.1%) | 12,744 (43.2%) | 6,575 (44.6%) |
| One Disease | 16,855 (33.9%) | 14,820 (34.0%) | 10,253 (34.7%) | 5,011 (34.0%) |
| Two or more diseases | 10,127 (20.3%) | 9,549 (21.9%) | 6,520 (22.1%) | 3,148 (21.4%) |
| **Types of PA** |  |  |  |  |
| None | 2,242 (4.5%) | 1,736 (4.0%) | 1,612 (5.5%) | 944 (6.4%) |
| Walk | 4,103 (8.2%) | 3,568 (8.2%) | 2,994 (10.1%) | 1,524 (10.3%) |
| Other Exercise | 2,363 (4.7%) | 1,993 (4.6%) | 1,440 (4.9%) | 687 (4.7%) |
| Sports | 268 (0.5%) | 217 (0.5%) | 126 (0.4%) | 90 (0.6%) |
| DIY | 2,043 (4.1%) | 2,094 (4.8%) | 1,755 (5.9%) | 1,385 (9.4%) |
| Two types of PA | 14,959 (30.1%) | 13,055 (30.0%) | 9,209 (31.2%) | 4,493 (30.5%) |
| Three types of PA | 14,699 (29.5%) | 12,873 (29.5%) | 7,941 (26.9%) | 3,701 (25.1%) |
| Four types of PA | 7,066 (14.2%) | 6,303 (14.5%) | 3,550 (12.0%) | 1,476 (10.0%) |
| All Types of PA | 2,024 (4.1%) | 1,735 (4.0%) | 890 (3.0%) | 434 (2.9%) |

values are frequency (percentage) unless otherwise stated. Accelerometer-derived variables were available only in a subset of participants. BMI = body mass index; PA = physical activity; MVPA = moderate-to-vigorous physical activity; MET = metabolic equivalent of task; DIY = do-it-yourself activity.

Compared with the very light OPA group, participants in the high OPA group were more likely to be male (62.3%), reside in more socioeconomically deprived areas (39.7% in the highest deprivation tertile), have lower educational attainment (15.8% with a college or university degree), and report lower household income (54.8% earning <£31,000 annually). High OPA was also associated with a greater prevalence of shift work (35.2% versus 5.8%) and longer working hours (>40 hours/week: 31.4% versus 26.1%).

Unhealthy lifestyle behaviors were more prevalent in the high OPA group, including current smoking (15.1%) and lower diet quality (35.2% in the least healthy tertile), alongside higher average alcohol consumption. Anthropometric and clinical measures showed modest gradients across OPA categories, with higher body mass index and systolic blood pressure observed in the high OPA group, while multimorbidity prevalence was similar across groups.

Accelerometer-derived measures demonstrated decreasing sedentary time and increasing MVPA across increasing OPA categories, with the high OPA group accumulating the greatest MVPA (mean 2823.7 MET-min/week). Estimated cardiorespiratory fitness (VOzmax) was similar across OPA categories. Leisure-time activity patterns were broadly comparable, although participants in the high OPA group were more likely to report engagement in do-it-yourself activities and less likely to report participation in three or more activity types.

### Association between Occupational Physical Activity and Incident Cardiovascular Disease

Over a median follow-up of 12.6 years (1,656,378 person-years), 10,465 incident cardiovascular disease (CVD) events occurred. Associations between OPA and incident CVD according to the two exposure classification methods are summarized in Figure 2, with full results from sequentially adjusted Cox proportional hazards models presented in Supplementary Tables S3-S7.

**Figure 2.**
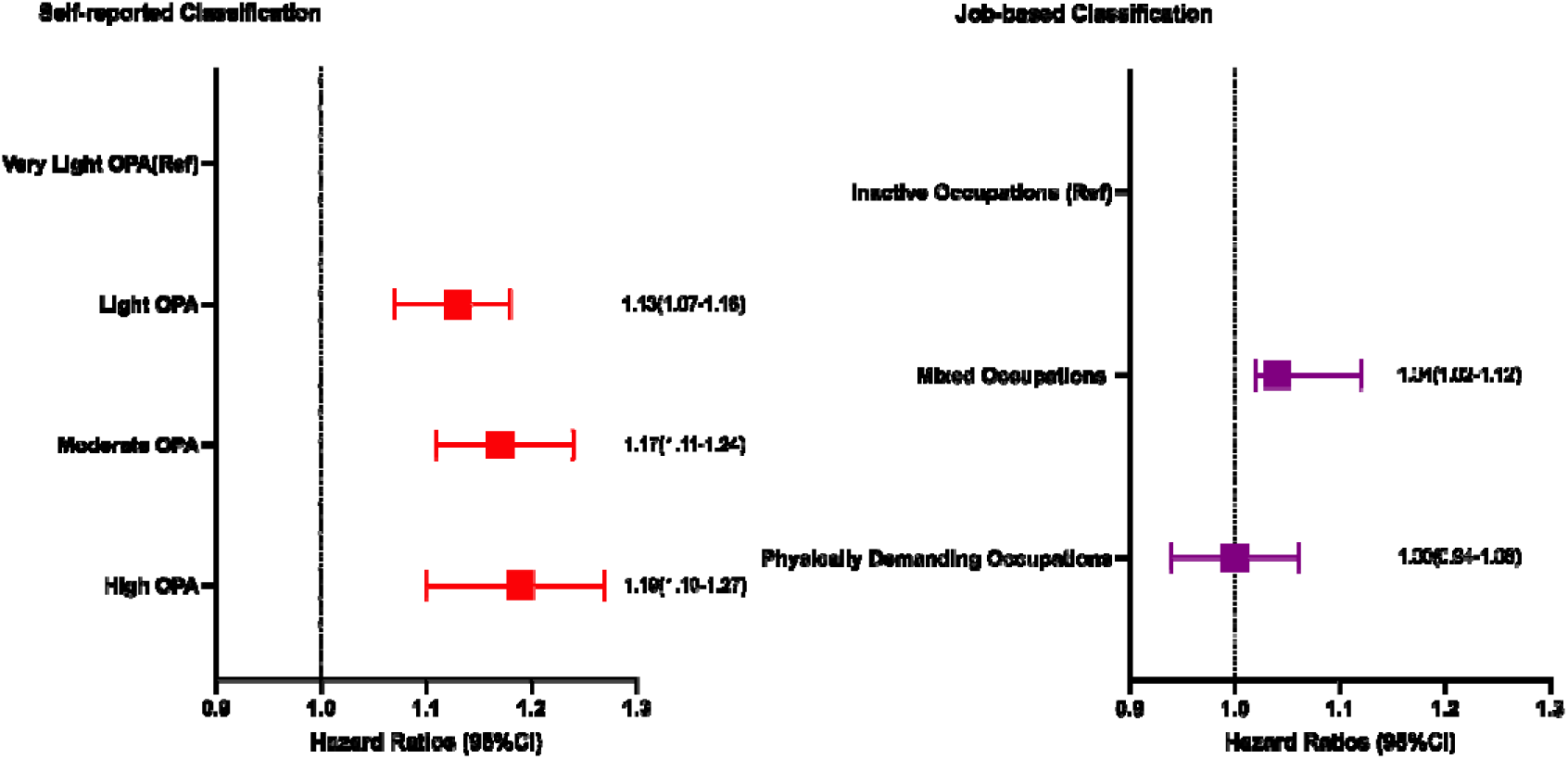
Association between occupational physical activity (OPA) and incident cardiovascular disease (CVD) according to two classification methods. Hazard ratios (HRs) and 95% confidence intervals (Cis) were estimated from fully adjusted Cox proportional hazards models. The left panel shows associations using individual-level self-reported OPA categories (very light, light, moderate, high), demonstrating a graded increase in CVD risk. The right panel shows group-based OPA categories derived from Standard Occupational Classification (SOC) codes (inactive, mixed, physically demanding), for which associations were attenuated after full adjustment. Models were adjusted for age, sex, ethnicity, socioeconomic status, education, income, smoking, alcohol intake, diet quality, body mass index, systolic blood pressure, diabetes, hypertension, multimorbidity, shift work, years in employment, weekly working hours, and leisure-time physical activity.

### Individual-level OPA classification

When OPA was classified using individual-level self-reported categories, higher OPA levels were associated with progressively increased CVD risk (Supplementary Table S3). In fully adjusted models, participants in the high OPA group had a 19% higher risk of CVD compared with those reporting very light OPA (hazard ratio [HR] 1.19, 95% confidence interval [CI] 1.11-1.27). Sequential adjustment resulted in marked attenuation of effect estimates, with the HR for high OPA declining from 1.60 in unadjusted analyses to 1.19 after full adjustment. These attenuation patterns across sequential models are illustrated in Supplementary Figure 1.

Further adjustment for psychosocial factors yielded similar estimates (HR 1.17, 95% CI 1.04-1.31), although analyses were restricted to participants with complete psychosocial data (Supplementary Table S4). Sex-stratified analyses demonstrated broadly comparable patterns in females and males, with similar degrees of attenuation following adjustment (Supplementary Table S5).

### Group-level OPA classification

When OPA was classified using group-level groupings derived from Standard Occupational Classification (SOC) codes, associations with CVD were substantially attenuated after adjustment (Supplementary Table S6). Although physically demanding occupations were associated with higher CVD risk in unadjusted models (HR 1.59, 95% CI 1.54-1.64), this association was fully attenuated after adjustment (HR 1.00, 95% CI 0.94-1.06). Sex-stratified analyses showed similar attenuation patterns in females and males, with no evidence of residual associations in fully adjusted models (Supplementary Table S7). The progressive attenuation and null associations observed after adjustment are visually summarized in Supplementary Figure 2.

Across both exposure definitions, sequential adjustment for demographic, socioeconomic, behavioral, clinical, occupational, and leisure-time PA factors resulted in progressive attenuation of effect estimates. However, only individual-level self-reported OPA retained a statistically significant association with incident CVD after full adjustment.

### Factors that Modify the Association between Occupational Physical Activity and Overall Cardiovascular Disease Incidence

Univariate regression analyses were conducted to examine the contribution of individual covariates to attenuation of the OPA-CVD association (Figure 3). The relative contribution of covariates differed by exposure classification, although several factors were consistent across both.

**Figure 3.**
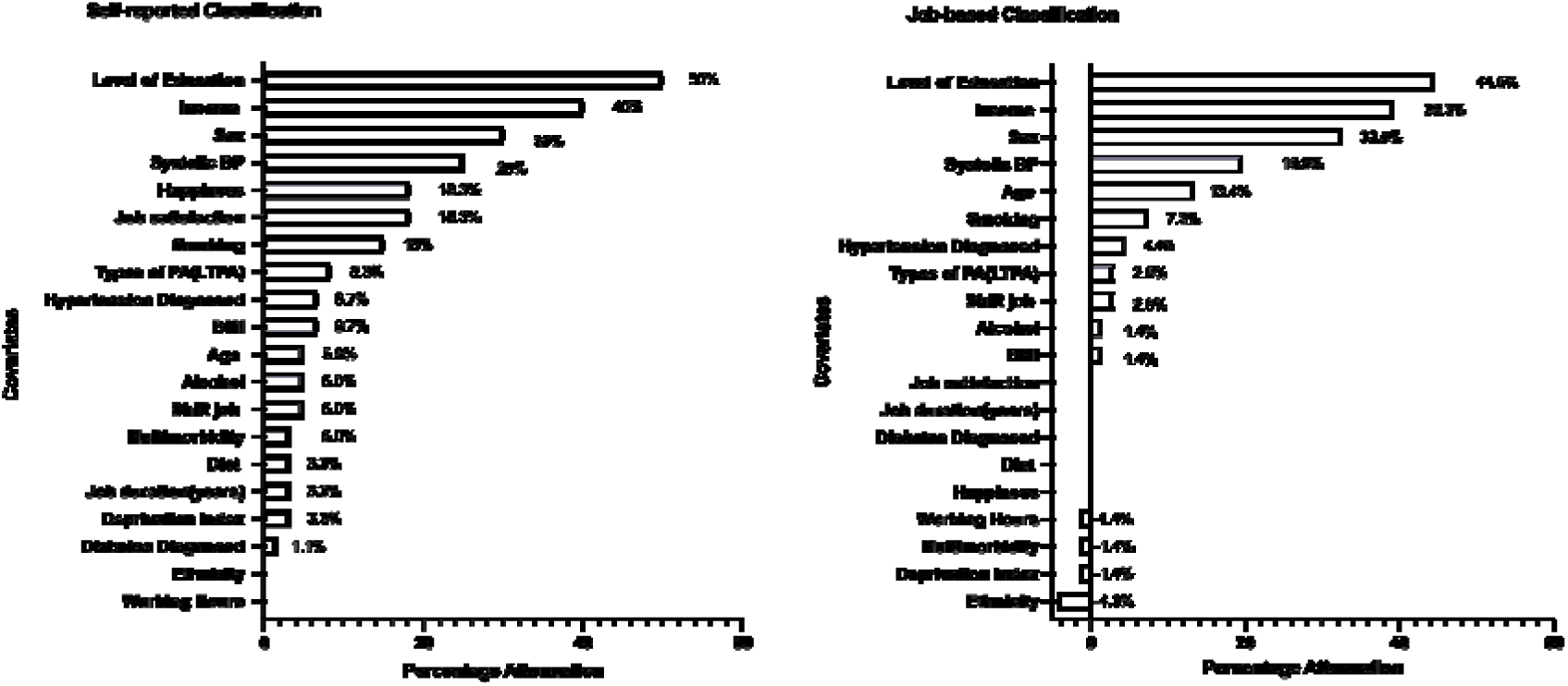
Factors modifying the association between occupational physical activity (OPA) and cardiovascular disease (CVD) incidence. Heller’s coefficient analysis of attenuation by covariates is shown for (A) Individual-level self-reported OPA classification and (B) Group-based classification using SOC codes. Bars represent the percentage attenuation in the hazard ratio for high OPA compared with the reference group after including each covariate individually in the model.

For individual-level self-reported OPA, education (50.0%), income (40.0%), and sex (30.0%) accounted for the largest proportion of attenuation, followed by systolic blood pressure (25.0%) and psychosocial factors, including happiness and job satisfaction (each 18.3%). Smaller contributions were observed for smoking, leisure-time PA, body mass index, and hypertension.

For the group-level classification, attenuation was primarily explained by education (44.5%), income (39.3%), and sex (32.5%), with additional contributions from systolic blood pressure (19.6%) and age (13.4%). Lifestyle, and occupational, contributed minimally, and psychosocial factors did not contribute to attenuation in the group-level classification.

## DISCUSSION

In this large prospective cohort study of 137,592 employed adults from the UK Biobank, the association between OPA and CVD incident was examined using alternative OPA classifications, alongside assessment of factors contributing to attenuation of the associations. The principal finding of this study show that the association between OPA and incident CVD differed substantially according to the level at which OPA was classified. Higher individual-level self-reported OPA was associated with increased CVD incidence, whereas no association was observed when OPA was assigned at the occupational-group level using SOC-based classifications. These findings suggest that conclusions regarding the PA paradox and CVD incidence are sensitive to how OPA is operationalised.

The divergence between the two approaches may reflect their different levels of exposure assignment. Individual self-reported OPA reflects the physical demands reported by each worker and may capture variation between workers within the same occupation. In contrast, the job-based approach assigns a common OPA category to members of the same SOC group according to the predominant physical demands observed within that group. Consequently, a worker whose reported activity differs from the prevailing pattern of their occupation may receive different OPA classifications under the two approaches. For example, a worker in an occupation predominantly classified as inactive may report substantial standing or walking, while an individual in a physically demanding occupational group may report relatively little PA because of differences in their specific role or duties. This suggest that exposure classification is not simply a methodological detail but may materially influence conclusions regarding the cardiovascular and potentially other effects of OPA [23].

These findings highlight the trade-off between feasibility and exposure specificity in occupational exposure assessment rather than suggesting that one classification approach is inherently superior. Group-level classifications have important practical advantages, as occupational information is routinely collected in many large datasets and can be applied at relatively low cost. However, their limited granularity may obscure substantial variation in physical demands within occupational groups. Individual-level assessment may better capture variation in workers’ reported physical demands, but requires more resource-intensive data collection. However, self-reported PA remains prone to reporting error, with studies showing limited agreement between self-reported and device-measured activity and a tendency towards overestimation [24, 25]. Future research should develop more nuanced exposure measures, such as Job Exposure Matrices linked to device-measured PA. However, several points need to be considered with this approach. Accurate domain-specific PA requires reliable information on working periods not currently detectable with device without additional information. Assuming a standard workday is inappropriate for many shift workers and physically demanding occupations [26] - groups well represented in the populations of interest for OPA research. In addition, in many large cohorts, including UK Biobank, device-measured PA was collected several years after baseline occupational and health data there creating a misalignment between variables [27]. Smaller scale, detailed studies using work diaries provide greater temporal resolution but are resource-intensive and often lack sufficient sample size to examine long-term health outcomes. The challenge for future research is therefore not simply to replace job-based classifications with individual-level or device-based measures, but to develop scalable approaches that combine occupational context with more precise, time-specific measures of PA.

Analyses of factors contributing to attenuation further highlight the importance of exposure classification. Across both approaches, socioeconomic position emerged as the dominant influence, with education and income accounting for the greatest attenuation. This is consistent with evidence linking lower socioeconomic position to physically demanding work, adverse health behaviours, reduced decision latitude and fewer resources to manage occupational stress [13, 28–30]. This pattern is consistent with effort-reward imbalance theory, whereby high demands coupled with low rewards may drive chronic stress and increased CVD risk [31]. Systolic blood pressure also explained a substantial proportion of the association, reflecting established links between OPA, hypertension and cardiovascular risk [21, 32]. However, the contribution of other factors differed according to the level at which OPA was classified. Psychosocial factors, including job satisfaction and happiness, contributed meaningfully only to associations based on individual self-reported OPA, whereas age emerged as an additional explanatory factor in group-level analyses. This is consistent with evidence that perceived work conditions and psychosocial stressors interact with physical workload to influence cardiovascular risk [33]. These findings suggest that the factors most apparent in explaining the OPA-CVD incidence association may also depend on how occupational exposure is operationalised. Additionally, the strong influence of socioeconomic factors regardless of how OPA was operationalised further highlights the importance of addressing structural determinants of occupational risk, including job quality, income inequality and working conditions. Workers in physically demanding roles may have limited capacity to engage in health-promoting behaviours outside work, reinforcing the importance of primary prevention within the workplace itself [19].

Strengths of the study include its large sample size, prospective design and comprehensive adjustment for potential confounders. A particular strength was the direct comparison of individual-level self-reported and occupational-group-level OPA classifications within the same cohort. This approach allowed assessment of whether conclusions regarding the OPA-CVD incidence association differed according to the level at which exposure was assigned, rather than assuming that alternative classifications were interchangeable. The cross-classified individual-level OPA measure also provided greater exposure granularity than single-item indicators.

Several limitations should be considered. As an observational study, residual confounding from unmeasured factors, including job strain, biomechanical load and workplace hazards, cannot be excluded. The two OPA classifications also involve different sources of potential exposure error. Individual-level self-reported OPA may be affected by reporting and classification error, whereas occupational-group-level SOC classifications assign a common exposure category according to predominant occupational demands and may therefore obscure within-occupation heterogeneity. Importantly, the use of self-reported OPA distributions to define SOC-level categories means that the two approaches are not fully independent and should not be interpreted as competing gold-standard measures. Rather, they represent different levels of OPA operationalisation. Exposure was assessed at a single time point, limiting assessment of changes in occupation or OPA over time and potentially contributing to exposure misclassification. Overall, the comparison of alternative OPA classifications highlights the importance of considering the level and approach of exposure assignment when interpreting associations between OPA and cardiovascular health.

## Conclusions

This study demonstrates that conclusions regarding the PA paradox and incident cardiovascular disease are sensitive to how OPA is classified. Individual-level self-reported OPA was associated with increased CVD incidence, whereas no association was observed when OPA was classified at the occupational-group level, highlighting exposure classification as a critical consideration in occupational epidemiology. Sociodemographic and health factors were key contributors to attenuation of associations regardless of classification, with psychosocial factors contributing primarily to individual-level self-reported OPA associations. These findings highlight exposure classification as a critical consideration when interpreting evidence on the PA paradox and the importance of addressing socioeconomic inequalities in OPA regardless of how OPA is operationalised.

## Data Availability

All data produced in the present study are available upon reasonable request to the authors

## Appendices: Supplementary Tables and Figures

**Supplementary Table 1.** Derivation of individual-level self-reported occupational physical activity (OPA) exposure groups.

| Manual/Physical Work | Standing/Walking work | Nine-group OPA category | Composite OPA Category (Primary analysis) |
| --- | --- | --- | --- |
| Low | Low | Low manual – Low Stand/walk | Very Light OPA |
| Low | Moderate | Low manual – Moderate Stand/walk | Light OPA |
| Low | High | Low manual – High Stand/walk | Light OPA |
| Moderate | Low | Moderate manual – Low Stand/walk | Light OPA |
| Moderate | Moderate | Moderate manual – Moderate Stand/walk | Moderate OPA |
| Moderate | High | Moderate manual – High Stand/walk | Moderate OPA |
| High | Low | High manual – Low Stand/walk | Moderate OPA |
| High | Moderate | High manual – Moderate Stand/walk | Moderate OPA |
| High | High | High manual – High Stand/walk | High OPA |

**Supplementary Table 2.** Group-level occupational physical activity (OPA) classification using SOC submajor groups.

| **Inactive Occupations** | **Mixed Occupations** | **Physically Demanding Occupations** |
| --- | --- | --- |
| Corporate Managers (11) | Managers and Proprietors in Agriculture and Services (12) | Skilled Agricultural Trades (51) |
| Science and Technology Professionals (21) | Health Professionals (22) | Skilled Metal and Electrical Trades (52) |
| Business and Public Professionals (24) | Teaching and Research Professionals (23) | Skilled Construction and Building Trades (53) |
| Culture, Media and Sports occupations (34) | Science and Technology Associate Professionals (31) | Textiles, Printing and Skilled Other Trades (54) |
| Business and Public Service Associate Professionals (35) | Health and Social Welfare Associate Professionals (32) | Process, Plant, and Machine Operatives (81) |
| Administrative Occupations (41) | Protective Service Occupations (33) | Elementary Trades, Plant and Storage Related Occupations (91) |
| Secretarial and Related Occupations (42) | Caring Personnel Service Occupations (61) | Elementary Administration and Service Occupations (92) |
| Customer Service Occupations (72) | Leisure and Other Personal Service Occupations (62) |  |
|  | Sales Occupations (71) |  |
|  | Transport and Mobile Machine Drivers and Operatives (82) |  |

Occupations were grouped using two-digit Standard Occupational Classification (SOC) sub-major groups derived from UK Biobank SOC codes. Group-based OPA categories were defined based on the distribution of self-reported OPA within each SOC group. SOC groups in which $\geq 70\%$ of participants reported very light or light OPA were classified as **inactive occupations**. SOC groups in which $\geq 60\%$ reported moderate or high OPA were classified as **physically demanding occupations**. SOC groups with a more even distribution across OPA categories were classified as **mixed occupations**. This dominance-based approach aligns with prior occupational epidemiology studies that classify jobs according to prevailing physical demands rather than assuming uniform exposure within occupations (9,13,25–27).

**Supplementary Table 3.**
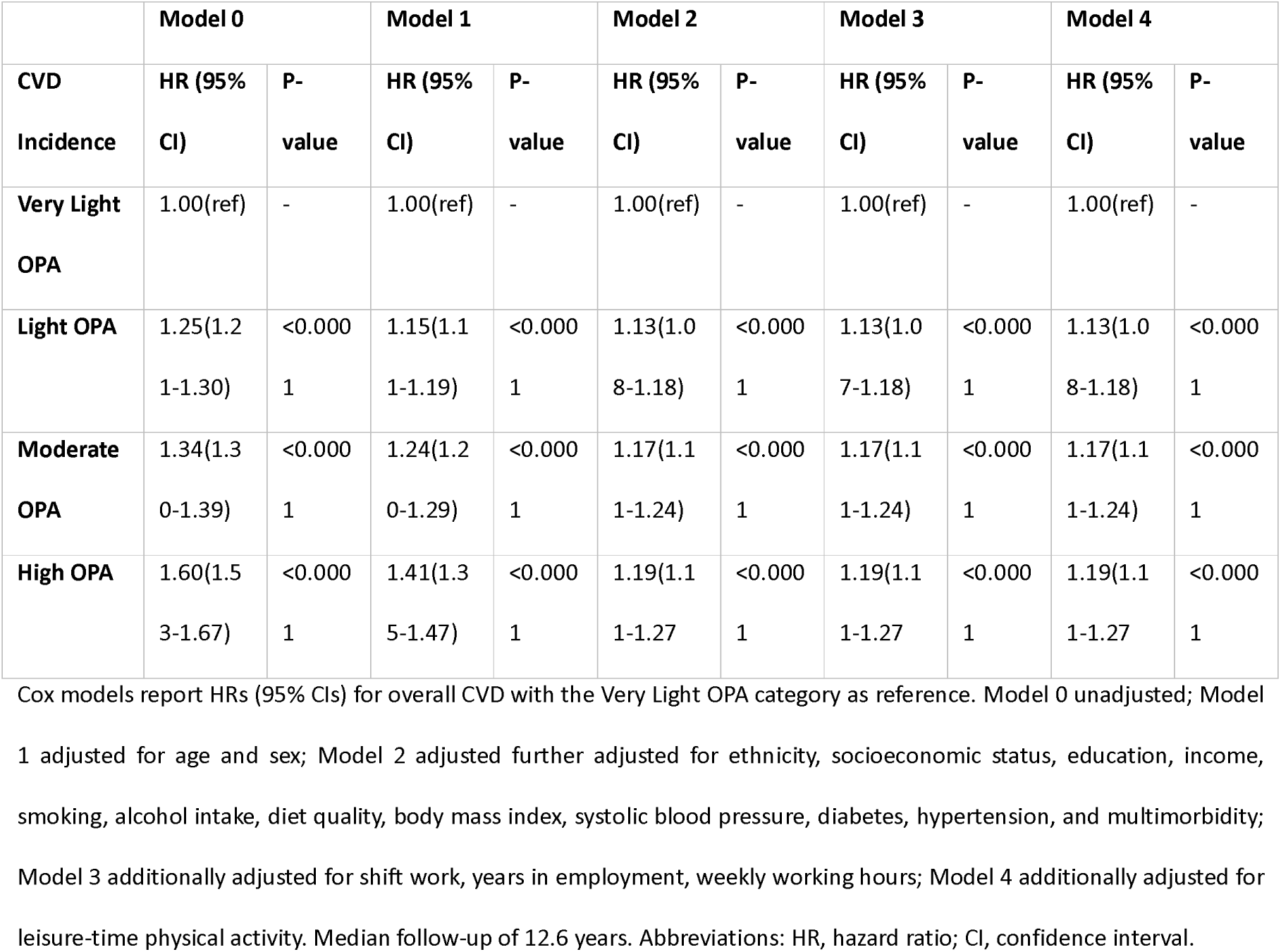
Association between OPA and CVD incidence: incremental adjustment models.

|  | **Model 0** |  | **Model 1** |  | **Model 2** |  | **Model 3** |  | **Model 4** |  |
| --- | --- | --- | --- | --- | --- | --- | --- | --- | --- | --- |
| **CVD Incidence** | **HR (95% CI)** | **P-value** | **HR (95% CI)** | **P-value** | **HR (95% CI)** | **P-value** | **HR (95% CI)** | **P-value** | **HR (95% CI)** | **P-value** |
| **Very Light OPA** | 1.00(ref) | - | 1.00(ref) | - | 1.00(ref) | - | 1.00(ref) | - | 1.00(ref) | - |
| **Light OPA** | 1.25(1.2<br>1-1.30) | <0.000<br>1 | 1.15(1.1<br>1-1.19) | <0.000<br>1 | 1.13(1.0<br>8-1.18) | <0.000<br>1 | 1.13(1.0<br>7-1.18) | <0.000<br>1 | 1.13(1.0<br>8-1.18) | <0.000<br>1 |
| **Moderate OPA** | 1.34(1.3<br>0-1.39) | <0.000<br>1 | 1.24(1.2<br>0-1.29) | <0.000<br>1 | 1.17(1.1<br>1-1.24) | <0.000<br>1 | 1.17(1.1<br>1-1.24) | <0.000<br>1 | 1.17(1.1<br>1-1.24) | <0.000<br>1 |
| **High OPA** | 1.60(1.5<br>3-1.67) | <0.000<br>1 | 1.41(1.3<br>5-1.47) | <0.000<br>1 | 1.19(1.1<br>1-1.27) | <0.000<br>1 | 1.19(1.1<br>1-1.27) | <0.000<br>1 | 1.19(1.1<br>1-1.27) | <0.000<br>1 |

Cox models report HRs (95% CIs) for overall CVD with the Very Light OPA category as reference. Model 0 unadjusted; Model 1 adjusted for age and sex; Model 2 adjusted further adjusted for ethnicity, socioeconomic status, education, income, smoking, alcohol intake, diet quality, body mass index, systolic blood pressure, diabetes, hypertension, and multimorbidity; Model 3 additionally adjusted for shift work, years in employment, weekly working hours; Model 4 additionally adjusted for leisure-time physical activity. Median follow-up of 12.6 years. Abbreviations: HR, hazard ratio; CI, confidence interval.

**Supplementary Table 4.** Sensitivity analysis: OPA and CVD incidence with psychosocial factors.

|  | **Model 5** |  |
| --- | --- | --- |
| **CVD Incidence** | **HR (95% CI)** | **P-value** |
| **Very Light OPA** | 1.00(ref) | - |
| **Light OPA** | 1.13(1.04-1.23) | 0 |
| **Moderate OPA** | 1.16(1.06-1.28) | 0 |
| **High OPA** | 1.17(1.04-1.31) | 0 |

Model 5 replicates Model 4 in the table above and further adjusts for job satisfaction and happiness. Analysis restricted participants with complete psychosocial data (n=51,778). Median follow-up of 12.6 years. Abbreviations: HR, hazard ratio; CI, confidence interval.

**Supplementary Table 5.** Sex-stratified association between OPA and CVD incidence.

| **Females** | **Model 0** |  | **Model 1** |  | **Model 4** |  |
| --- | --- | --- | --- | --- | --- | --- |
| **CVD Incidence** | **HR (95% CI)** | **P-value** | **HR (95% CI)** | **P-value** | **HR (95% CI)** | **P-value** |
| **Very Light OPA** | 1.00(ref) | - | 1.00(ref) | - | 1.00(ref) | - |
| **Light OPA** | 1.19(1.13-1.27) | <0.0001 | 1.15(1.09-1.22) | <0.0001 | 1.12(1.03-1.22) | 0.01 |
| **Moderate OPA** | 1.35(1.27-1.43) | <0.0001 | 1.24(1.17-1.32) | <0.0001 | 1.13(1.04-1.24) | 0.01 |
| **High OPA** | 1.56(1.44-1.68) | <0.0001 | 1.53(1.42-1.66) | <0.0001 | 1.23(1.09-1.40) | 0.001 |
| **Males** |  |  |  |  |  |  |
| **CVD Incidence** | HR (95% CI) | P-value | HR (95% CI) | P-value | HR (95% CI) | P-value |
| **Very Light OPA** | 1.00(ref) | - | 1.00(ref) | - |  |  |
| **Light OPA** | 1.22(1.17-1.27) | <0.0001 | 1.14(1.09-1.19) | <0.0001 | 1.14(1.07-1.21) | <0.0001 |
| **Moderate OPA** | 1.37(1.31-1.43) | <0.0001 | 1.24(1.19-1.30) | <0.0001 | 1.21(1.12-1.29) | <0.0001 |
| **High OPA** | 1.38(1.31-1.45) | <0.0001 | 1.36(1.29-1.43) | <0.0001 | 1.19(1.09-1.29) | <0.0001 |

Hazard ratios (HRs) with 95% confidence intervals (CIs) estimated from Cox proportional hazards models, using the very light OPA group as reference. Results are presented separately for females and males.
Median follow-up of 12.6 years. Abbreviations: HR, hazard ratio; CI, confidence interval.

**Supplementary Table 6.** Association between group-level classification and CVD incidence: incremental adjustment models.

|  | Model 0 |  | Model 1 |  | Model 2 |  | Model 3 |  | Model 4 |  |
| --- | --- | --- | --- | --- | --- | --- | --- | --- | --- | --- |
| CVD Incidence | HR (95% CI) | P-value | HR (95% CI) | P-value | HR (95% CI) | P-value | HR (95% CI) | P-value | HR (95% CI) | P-value |
| Inactive Occupations | 1.00(ref) | - | 1.00(ref) | - | 1.00(ref) | - | 1.00(ref) | - | 1.00(ref) | - |
| Mixed Occupations | 1.06(1.03-1.09) | <0.0001 | 1.11(1.08-1.14) | <0.0001 | 1.07(1.03-1.11) | 0.001 | 1.07(1.02-1.12) | 0.004 | 1.07(1.02-1.12) | 0.004 |
| Physically Demanding Occupations | 1.59(1.54-1.64) | <0.0001 | 1.27(1.23-1.31) | <0.0001 | 1.02(0.97-1.08) | 0.38 | 1.00(0.94-1.06) | 0.95 | 1.00(0.94-1.06) | 0.95 |

Cox models report HRs (95% CIs) for overall CVD with the inactive job category as reference. Model 0 unadjusted; Model 1 adjusted for age and sex; Model 2 adjusted further adjusted for ethnicity, socioeconomic status, education, income, smoking, alcohol intake, diet quality, body mass index, systolic blood pressure, diabetes, hypertension, and multimorbidity; Model 3 additionally adjusted for shift work, years in employment, weekly working hours; Model 4 additionally adjusted for leisure-time physical activity. Median follow-up of 12.6 years. Abbreviations: HR, hazard ratio; CI, confidence interval.

**Supplementary Table 7.** Sex-stratified association between group-level classification and CVD incidence.

| Females | Model 0 |  | Model 1 |  | Model 4 |  |
| --- | --- | --- | --- | --- | --- | --- |
| CVD Incidence | HR (95% CI) | P-value | HR (95% CI) | P-value | HR (95% CI) | P-value |
| Inactive Occupations | 1.00(ref) | - | 1.00(ref) | - | 1.00(ref) | - |
| **Mixed Occupations** | 1.13(1.08-1.17) | <0.0001 | 1.07(1.02-1.11) | 0.002 | 1.07(1.00-1.15) | 0.07 |
| **Physically Demanding Occupations** | 1.60(1.49-1.71) | <0.0001 | 1.41(1.31-1.51) | <0.0001 | 0.94(0.82-1.08) | 0.36 |
| **Males** |  |  |  |  |  |  |
| **CVD Incidence** | HR (95% CI) | P-value | HR (95% CI) | P-value | HR (95% CI) | P-value |
| **Inactive Occupations** | 1.00(ref) | - | 1.00(ref) | - |  |  |
| **Mixed Occupations** | 1.22(1.17-1.26) | <0.0001 | 1.15(1.11-1.20) | <0.0001 | 1.08(1.02-1.15) | 0.01 |
| **Physically Demanding Occupations** | 1.30(1.26-1.35) | <0.0001 | 1.24(1.20-1.29) | <0.0001 | 1.03(0.96-1.1) | 0.43 |

Hazard ratios (HRs) with 95% confidence intervals (CIs) estimated from Cox proportional hazards models, using the very light OPA group as reference. Results are presented separately for females and males.
Median follow-up of 12.6 years. Abbreviations: HR, hazard ratio; CI, confidence interval.

**Supplementary Figure 1:**
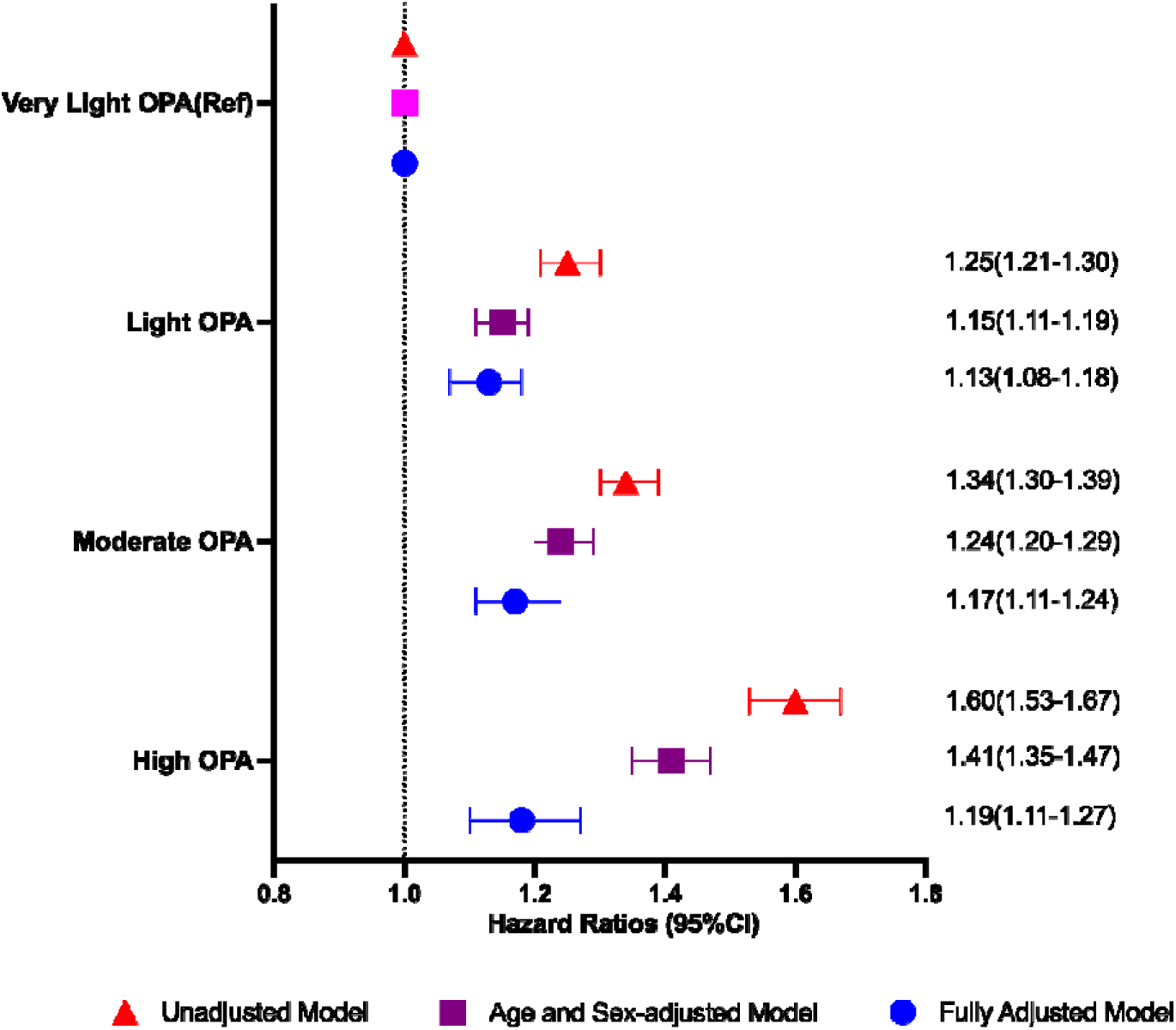
Association between Occupational Physical Activity (OPA) and cardiovascular disease (CVD) Incidence. Data presented as hazard ratio and 95% CI estimates from Cox proportional hazards models, with the very light OPA group as the reference. Three models are presented: Model 0 (unadjusted); Model 1 (adjusted for age and sex); Model 4 which was fully adjusted for age, sex, ethnicity, socioeconomic status, education, income, smoking, alcohol intake, diet quality, body mass index, systolic blood pressure, diabetes, hypertension, multimorbidity, shiftwork, years in employment, weekly working hours, and leisure-time physical activity.

**Supplementary Figure 2:**
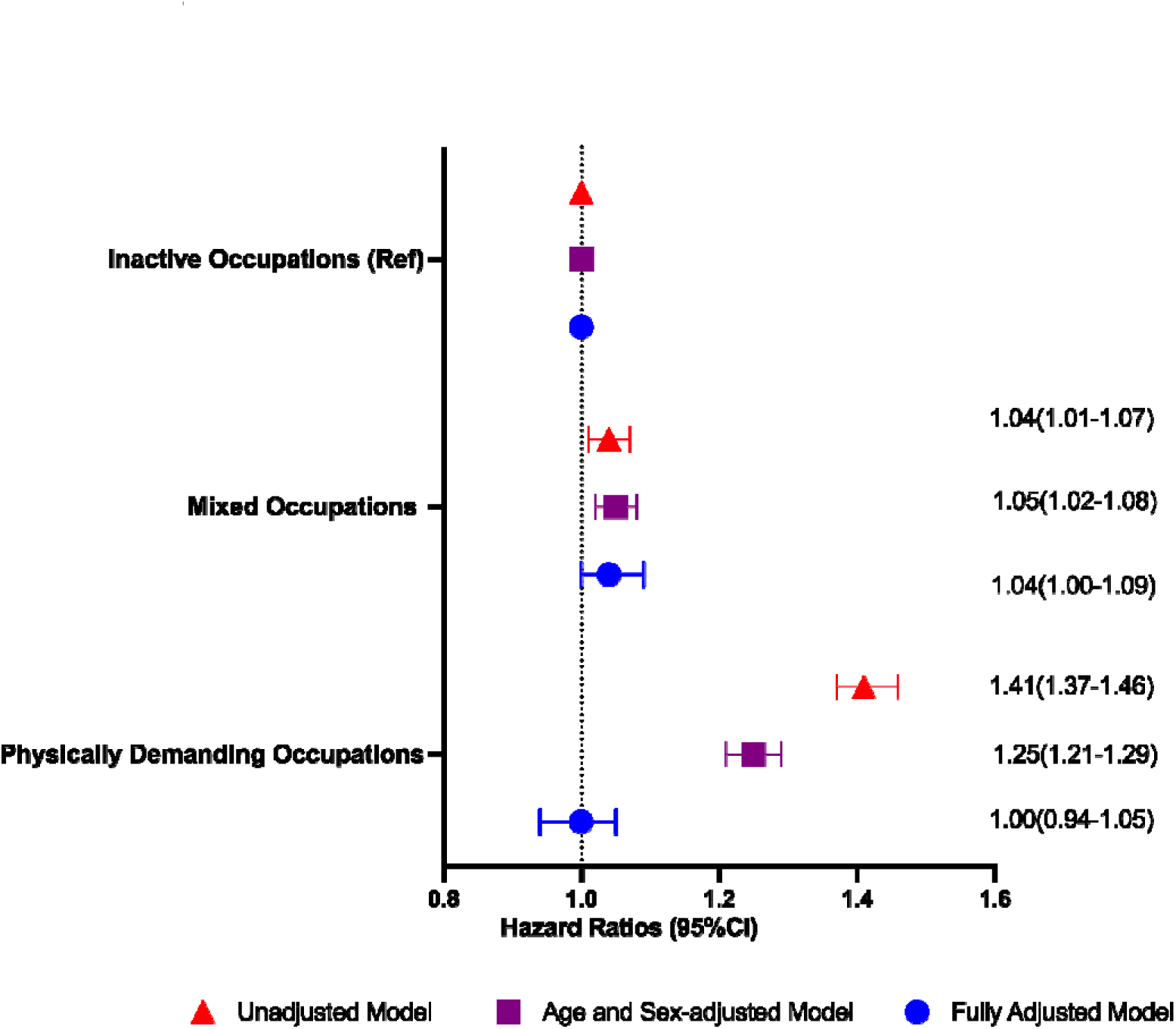
Association between group-level OPA classification and CVD Incidence. Data presented as hazard ratio and 95% CI estimates from Cox proportional hazards models, with inactive occupations as the reference. Three models are presented: Model 0 (unadjusted); Model 1 (adjusted for age and sex); Model 4 which was fully adjusted for age, sex, ethnicity, socioeconomic status, education, income, smoking, alcohol intake, diet quality, body mass index, systolic blood pressure, diabetes, hypertension, mu Hi morbidity, shift work, years in employment, weekly working hours, and leisure-time physical activity.

